# Dietary pattern and diversity analysis using ‘DietR’ package in R

**DOI:** 10.1101/2023.07.07.23292390

**Authors:** Rie Sadohara, David Jacobs, Mark A. Pereira, Abigail J. Johnson

## Abstract

There are scarce resources available for analyzing 24-hour dietary records. Here we introduce DietR, a set of functions written in R for the analysis of 24-hour dietary recall or records data, collected with either the Automated Self-Administered 24-hour (ASA24) dietary assessment tool or two-day data from the National Health and Nutrition Examination Survey (NHANES). The R functions are intended for food and nutrition researchers who are not computational experts. DietR provides users with functions to (1) clean dietary data; (2) analyze 24-hour dietary intakes in relation to other study-specific metadata variables; (3) visualize percentages of calorie intake from macronutrients; (4) perform principal component analysis (PCA) or *k*-means to group participants by similar dietary patterns; (5) generate foodtrees based on the hierarchical information of food items consumed; (6) perform principal coordinate analysis (PCoA) taking food classification information into account; (7) and calculate diversity metrics for overall diet and specific food groups. DietR includes a set of tutorials available on a website (https://computational-nutrition-lab.github.io/DietR/), which are designed to be self-paced study materials. DietR enables users to visualize dietary data and conduct data-driven dietary pattern analyses using R to answer research questions regarding diet. As a demonstration of DietR, we applied DietR to a set of created 24-hour dietary records data to demonstrate the basic functions of the package. We also applied DietR to a subset of 24-hour recall data from NHANES to demonstrate analyses using dietary diversity metrics. We present the results of this example NHANES analysis comparing legume diversity with waist circumference.

## Introduction

Data-driven dietary pattern analysis is increasingly being applied to large datasets of 24-hour dietary recall data. Efforts like the proposed Nutrition for Precision Health, powered by the All of Us research program (https://allofus.nih.gov/), are expected to collect 24-hour dietary recall data which will be made publicly available. While methods exist to calculate dietary patterns from food frequency questionnaire data (1), using 24-hour recall data poses unique challenges. As more and larger datasets are published, there is also a need for reproducibility and the ability to combine datasets. A majority of the published code for the analysis of 24-hour dietary data is written for proprietary softwares, posing issues for open data science practices. For example, code to analyze Automated Self-Administered 24-hour (ASA24) dietary assessment tool is available only for SAS (SAS Institute Inc., Cary, NC). R is an open-source platform with countless packages and public resources that can be tailored to one’s statistical analysis and visualization needs (2). Therefore, using R for dietary data analysis will increase accessibility and reproducibility, as R is available without a license fee. Despite the benefits to researchers gained by using R, resources are scarce for analyzing 24-hour recall data using R; therefore, we present a dietary analysis toolkit, DietR, currently supporting ASA24 and NHANES data. This package contains R functions for data loading and preparation, exploratory data overview, clustering analysis, food tree generation, ordination, and diversity metrics (**Table 1**) with an aim to facilitate data-driven dietary pattern analysis by beginner-intermediate R users. The dependency R packages of DietR are listed in **Supplementary Table 1**. We applied DietR to a set of created 24-hour dietary records data to demonstrate the basic functions of the package. We also applied DietR to a subset of 2-day recall data from NHANES to demonstrate analyses with dietary diversity metrics. We present the results of this NHANES analysis comparing legume diversity with waist circumference.

**Table 1.** DietR package features.

| Functionality | Description |
| --- | --- |
| Load raw data | Load ASA24 data (.csv) or NHANES (.XPT) to R, compute totals from items data, and compute means across record collection days. |
| Clean data | Remove incomplete data, apply filters to participants and to dietary data according to specified conditions and/or ASA24 data cleaning guidelines. |
| Summarize data | Generate basic statistics of variables in the totals data, and generate boxplots and scatterplots of variables of interest. |
| Generate variables to group participants | Generate variables based on their demographic, health, or other data to group them. e.g. GLU-index by their glucose tolerance test results, or Edu variable based on their educational background. |
| Visualize %energy from macronutrients | Compute % of energy intake from carbohydrate, protein, and total fat, and visualize the proportions in barplots. |
| Prepare for clustering | Keep complete rows and remove variables with zero variance and highly correlated variables. |
| Perform clustering analyses | Perform PCA<sup>a</sup> or *k*-means analyses with nutrient or food category data and generate biplots and barplots of variables contributing to PCs. |
| Generate foodtrees | Generate foodtrees comprised of consumed food items that are grouped according to their similarity to one another. |
| Visualize foodtrees | Visualize foodtrees in a circular plot at a desired classification depth. |
| Perform ordination | Compute distances between participants and perform clustering analyses such as PCoA<sup>b</sup> with the hierarchical information of foods taken into consideration. Test differences between groups by permanova. |
| Visualize ordination results | Generate biplots with participants color-coded by a variable of interest. |
| Inspect food items correlated with ordination axes | Show correlation coefficients and q-values for food items and selected ordination axes. |
| Compute diversity of all or specific food groups | Compute alpha-diversity of food items consumed and generate a categorical variable based on the diversity. |

a: principal component analysis; b: principal coordinate analysis.

## Methods

### Demonstration data set

The DietR package comes with example scripts to prepare, visualize, and analyze dietary data as dietary patterns and dietary diversity (**Figure 1**). For ASA24 data analysis demonstrations, we created a dataset of dietary records (referred to as VVKAJ). VVKAJ is a hypothetical study with three days of dietary records from 17 fictitious participants following one of five diets: Vegetarian, Vegan, Keto, American, and Japanese. Menus for these demonstration data were selected from online blogs and recipes, with a goal of creating meal patterns that would differ by macronutrient profile and food selection.

**Figure 1.**
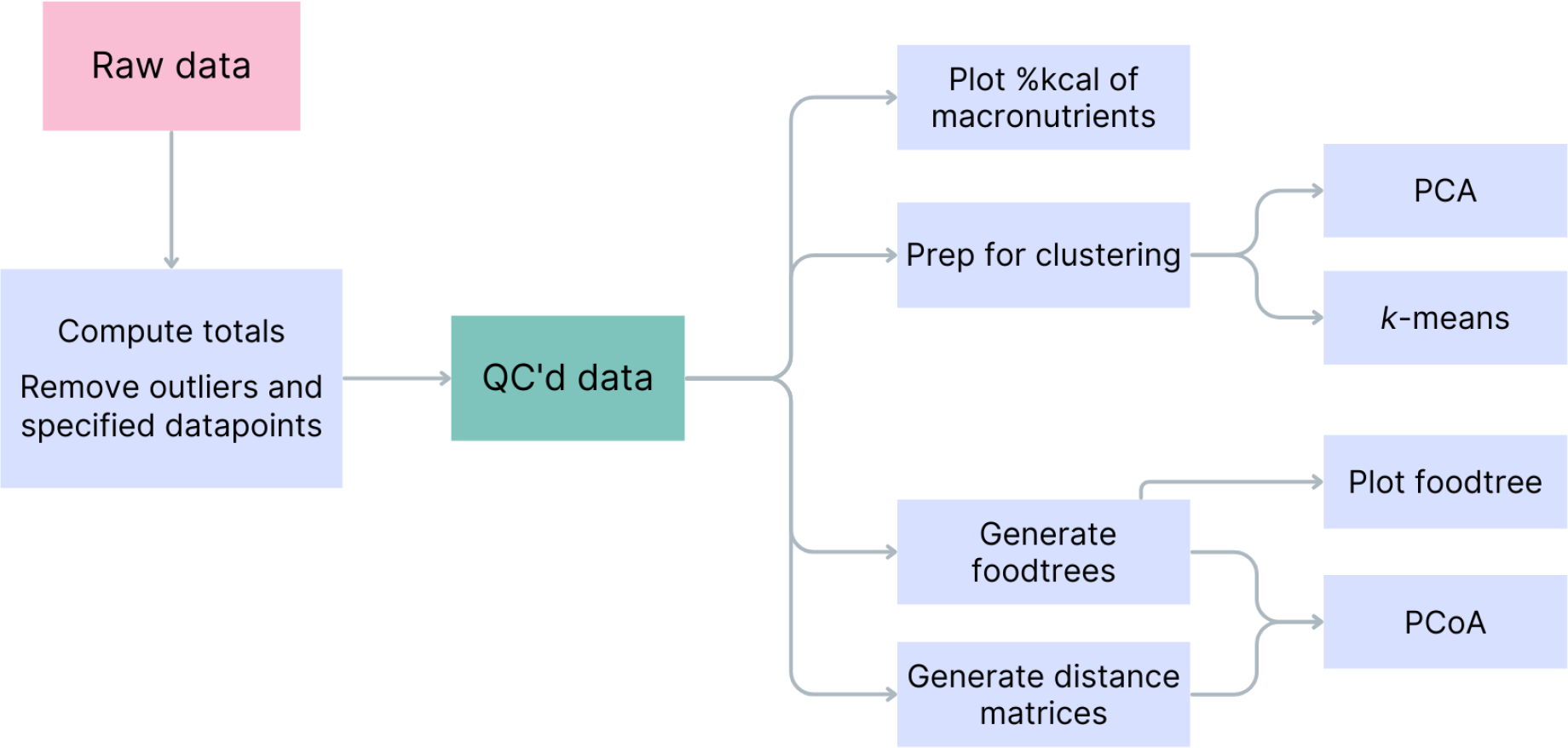
Flow of dietary data analyses using the DietR package.

#### Data preparation

We applied functions from the DietR package to load, filter outliers and missing data, compute total food intake for each participant and day, compute means of food intake across days, and quality-control procedures for further analyses. The quality control of mean totals data can be done according to the ASA24 data outliers guideline (3), or a threshold for a variable set by the user.

#### Clustering analysis

To run clustering analysis, we first removed highly correlated variables. We filtered variables with zero variance and highly correlated variables at a specified threshold of correlation coefficient (r>0.75). We used the set of remaining variables with no missing data and non-zero variance as an input for *k*-means and principal component analyses (PCA). The DietR package uses functions from the ggplot2 (4) and factoextra (5) packages to visualize those clustering results in the form of bi-plots, scatter plots, and bar plots, with the individuals color-coded by their groups.

#### Tree-based analysis

The DietR package includes functions to generate foodtrees, where food items are hierarchically grouped (6), such that nine categories of foods branch into more detailed classifications. By taking into account the food items’ phenetic relationships, the participant’s dietary patterns can be analyzed not only by the food items they consumed, but also by the similarity or food group of the foods they consumed. Tree visualizations can be generated with color-coded food groups to visually explore the participant’s overall intake by using the ggtree package (7). We used the phyloseq and vegan packages (8,9) to perform tree-based Principal Coordinate Analysis (PCoA) with weighted or unweighted UniFrac distances (10) between participants. After checking for differences in beta-dispersion, we tested for between-group dietary separation using a permutation-based statistical test (PERMANOVA, 5,000 permutations) and the pairwiseAdonis function (11).

### NHANES dataset

#### Record filtering

Food items records were obtained from the NHANES 2015-16, which was cleaned by selecting participants with complete data entry, who were 18 years or older, and reported more than one food item per day (n=4,402). Population-wise adjustment using survey weights was omitted here for the sake of simple demonstration of the utility of this package. A total intake was calculated by summing food intakes for each participant for each day of the two data collection days. The two-day means of totals were calculated for each participant. To remove potential outlier records, the two-day means of totals were cleaned in accordance with the General Guidelines for Reviewing & Cleaning Data by ASA24 (3); namely, total calories (KCAL) <650 or >5700, total protein (PROT, g) <25 or >240, total fat (TFAT, g) <25 or >230, vitamin C (VC, mg) <5 or >400 for males, and total calories (KCAL) <600 or >4400, total protein (PROT, g) <10 or >180, total fat (TFAT, g) <15 or >185, vitamin C (VC, mg) <5 or >350 for females. Criteria for beta-carotene (BCAR, <15 or >8200 μg for males and <15 or >7100 μg for females) was not used because the records with beta-carotene intake outside the criteria did not seem to be outliers. As a result, 4,164 individuals were retained. Furthermore, the individuals with complete body mass index (BMI) and waist circumference data were retained (n=3,998). The sociodemographic variables, Age, Gender, Ethnicity, Income, and Education were included in the NHANES 2015-16 totals data and were used as covariates. The ratio of family income to poverty (Family IPR) was used as the income variable, and 356 records with missing data in this variable were excluded (n=3,642). The other demographic variables did not have missing data.

#### Grouping by the diversity in nuts/seeds/legumes consumption

Food items from NHANES with FNDDS (Food and Nutrient Database for Dietary Studies) foodcodes between 40000000 - 49999999, which are nuts, seeds, and legumes, were selected from the QC-ed food items of the 3,642 participants. An individual food consumption (IFC) table was generated with nuts/seeds/legumes consumed during the 2 recorded days for each participant, and the average consumption (g/day) was computed. One participant recorded with an excessive amount (1,500 g/day) of nuts/seeds/legumes consumption was considered to be an outlier and was excluded. All other records had nuts/seeds/legumes consumption < 1,000 g/day. As a result, 3,641 records were included. From the IFC table, Shannon’s diversity index for nuts/seeds/legumes was calculated for each participant. The data were divided into groups for analysis based on the diversity in the nuts/seeds/legumes items they consumed. Those who consumed zero nuts/seeds/legumes foods in the two days (n=1,819) were grouped as DivNA. Those who consumed only one nuts/seeds/legumes food (n=1,105) had a diversity index of 0 (only one food, thus no diversity), and they were labeled as Div0. Those who consumed more than one nuts/seeds/legumes food (n=717) with a diversity index greater than 0 were split into the lower and upper halves, which were named Div1 and Div2, respectively. Div1 group’s diversity index ranged from 0.027 to 0.66, and Div2 group’s diversity index ranged from 0.66 to 1.95 (**Table 2**).

**Table 2.**
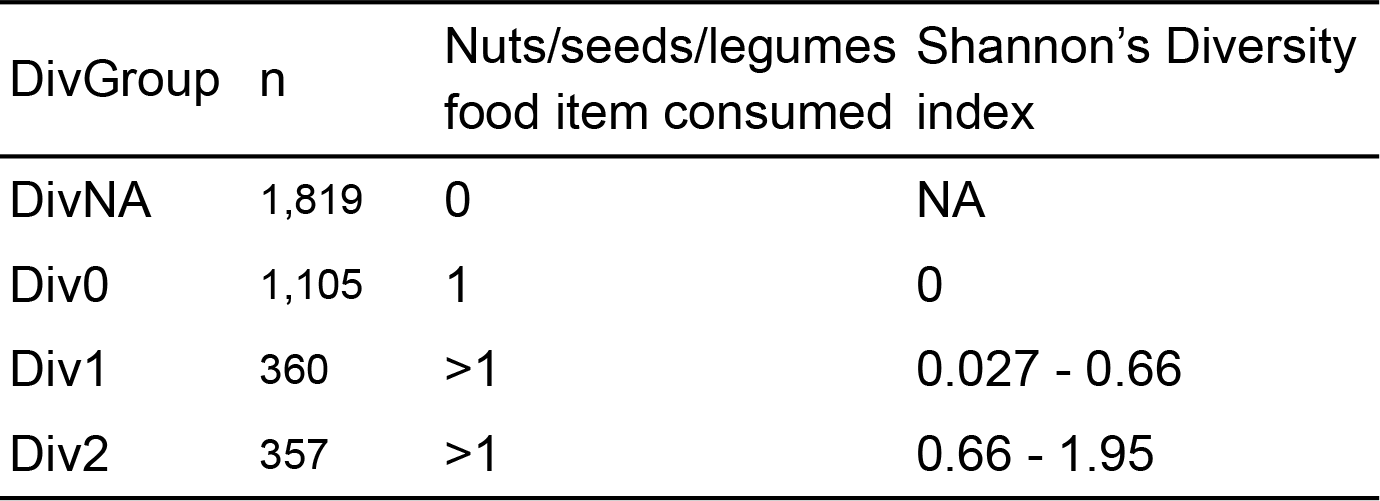
Nuts/seeds/legumes diversity groups with complete BMI and waist circumference measurements.

#### Sociodemographic and nutrients variables

The participants were grouped into three age groups: 18-39, 40-59, and 60 years or older; three income groups: Family income to poverty ratio (IPR) < 1.85, 1.85-2.99, and ≥3.00; and three education groups: < high school, high school graduate or some college, and college graduate or above. The chisq.test function in base R was used to test the deviation of expected and observed ratios between the categories in each of the sociodemographic variables and the DivGroups (**Table 3**). The means of the quantity of protein foods excluding legumes, legumes, total protein foods, the percentages of legumes (if consumed) in total protein food consumption, total vegetable intake except legumes, and total fruit intake were computed for each of the DivGroups. Macronutrients (carbohydrate, protein, and total fat), dietary fiber, total saturated fat, total unsaturated fat (mono- and poly-unsaturated fats), alcohol, and added sugar intakes were adjusted per 2,000 kcal (**Table 4**).

**Table 3.** Sociodemographic features of the DivGroups.

|  | DivNA<br>(n=1819) | Div0<br>(n=1105) | Div1<br>(n=360) | Div2<br>(n=357) |  |
| --- | --- | --- | --- | --- | --- |
|  | ----- % ----- |  |  |  | *p*-Chi sq. |
| Gender |  |  |  |  | 0.283 |
| Male | 49 | 46 | 46 | 45 |  |
| Female | 51 | 54 | 54 | 55 |  |
| Age |  |  |  |  | 0.009 |
| 18-39 | 39 | 34 | 32 | 32 |  |
| 40-59 | 31 | 33 | 37 | 35 |  |
| 60+ | 30 | 33 | 31 | 34 |  |
| Race/Ethnicity |  |  |  |  | < 0.0001 |
| Mexican Am. & Hispanic | 27 | 35 | 28 | 30 |  |
| NH<sup>a</sup> White | 37 | 36 | 38 | 36 |  |
| NH Black | 26 | 17 | 13 | 9 |  |
| NH Asian | 6 | 8 | 19 | 23 |  |
| Other race | 4 | 4 | 3 | 2 |  |
| Family IPR<sup>b</sup> |  |  |  |  | < 0.0001 |
| <1.85 | 49 | 44 | 33 | 32 |  |
| 1.85-2.99 | 21 | 20 | 22 | 18 |  |
| ≥ 3.00 | 29 | 35 | 45 | 50 |  |
| Education |  |  |  |  | < 0.0001 |
| < HS<sup>c</sup> | 21 | 21 | 13 | 13 |  |
| HS grad or some collage | 59 | 52 | 46 | 40 |  |
| Collage grad or above | 20 | 28 | 41 | 47 |  |

a: Non-Hispanic; b: family-wise income to poverty ratio; c: high school.

**Table 4.** Dietary features of the DivGroups, consumption per day, means and (SD).

| Variable | Unit | DivNA | Div0 | Div1 | Div2 | p-ANOVA<sup>a</sup> |
| --- | --- | --- | --- | --- | --- | --- |
| **Food intake** |  |  |  |  |  |  |
| Legume intake | oz. | 0.2 (0.6) | 0.7 (1.1) | 1.1 (1.5) | 1.4 (1.9) | <0.0001 |
| PF<sup>b</sup> + legumes | oz. | 5.8 (3.4) | 7.0 (3.4) | 8.2 (3.8) | 8.7 (4.3) | <0.0001 |
| Legumes in total PF intake | % | - | 11.1 (16.2) | 15 (18.8) | 17.4 (20.8) | <0.0001<sup>c</sup> |
| Total fruit | cup | 0.8 (0.9) | 1.0 (1.0) | 1.2 (1.2) | 1.4 (1.4) | <0.0001 |
| Total vegetables excluding legumes | cup | 1.4 (0.9) | 1.5 (1.0) | 1.8 (1.0) | 1.7 (1.0) | <0.0001 |
| Nuts/seeds/legumes consumption | g/day | 0 (0) | 49 (57) | 104 (102) | 131 (107) | <0.0001<sup>c</sup> |
| Total number of items reported | ct. | 14 (5) | 16 (5) | 18 (5) | 19 (5) | <0.0001 |
| Total energy | kcal | 1960 (714) | 2031 (694) | 2136 (691) | 2192 (731) | <0.0001 |
| **Nutrient intake<sup>d</sup>** |  |  |  |  |  |  |
| Carbohydrate | g | 241 (45) | 238 (42) | 239 (45) | 239 (45) | 0.476 |
| Protein | g | 79 (22) | 82 (21) | 82 (20) | 82 (19) | <0.001 |
| Dietary fiber | g | 15 (6) | 18 (7) | 22 (9) | 24 (9) | <0.0001 |
| Total fat | g | 77 (16) | 78 (16) | 78 (17) | 80 (18) | <0.01 |
| Total saturated fat | g | 26 (7) | 25 (7) | 23 (7) | 23 (7) | <0.0001 |
| Total monounsaturated fat | g | 27 (6) | 28 (7) | 28 (8) | 30 (9) | <0.0001 |
| Total polyunsaturated fat | g | 17 (6) | 18 (6) | 20 (7) | 20 (6) | <0.0001 |
| Total unsaturated fat | g | 44 (11) | 46 (11) | 48 (12) | 50 (14) | <0.0001 |
| Alcohol | g | 7 (17) | 6 (14) | 6 (14) | 5 (12) | 0.095 |
| Added sugars | tsp. | 17 (11) | 14 (8) | 12 (8) | 11 (7) | <0.0001 |

a: *p*-value for ANOVA test for DivGroups; b: protein foods; c: tested with Div0, Div1, and Div2, excluding DivNA; d: adjusted per 2,000 kcal.

#### Analysis of covariance for body measurements

To analyze body measurements (BMI and waist circumference) for each DivGroup, the car package (12) in R was used to run type III analysis of covariance (ANCOVA) model with DivGroup as the treatment effect and Age, Gender, Ethnicity, Income, Education, and kcal intake as covariates. The estimated marginal means (emmeans) were used to statistically test for associations between waist circumference or BMI and the DivGroups and conduct pairwise comparisons between DivGroups by using the emmeans package (13).

## Results

### Demonstration data highlights the utility of DietR

Exploration of the demonstration dataset with DietR showed expected variability in macronutrient content (**Figure 2A**) and separation in principal coordinate space (**Figure 2B**). A food tree was created from the data (**Figure 2C**), and this was used to show separation in a tree-based PCoA (*p*-value=2×10^-4^, PERMANOVA, alpha=0.05) (**Figure 2D**).

**Figure 2.**
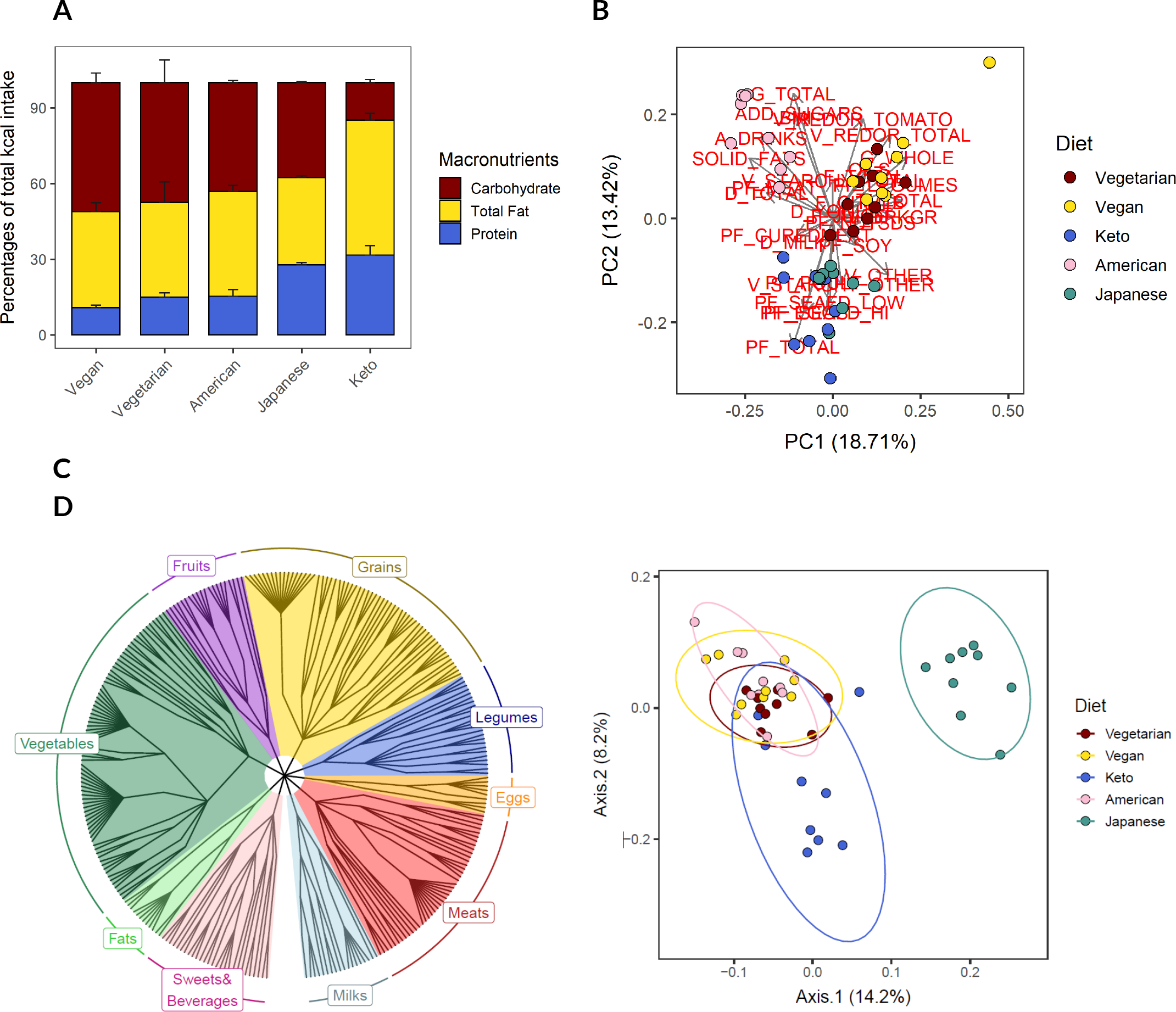
Charts that can be generated by analyzing the VVKAJ (Vegetarian, Vegan, Keto, American, and Japanese diets) dataset using the functions in the DietR package. **A**: Percentages of total calorie intake by macronutrients for various diet groups. **B**: Biplot of Principal Component Analysis (PCA) carried out with the food consumption data by food category of the participants. **C**: Foodtree of the reported foods by the participants organized by the first level of food categories: Milks, Meats, Eggs, Legumes, Grains, Fruits, Vegetables, Fats, and Sweets&Bevarages. **D**: Biplot of Principal Coordinate Analysis (PCoA) carried out with weighted unifrac distance of reported food items and their consumption amounts by the participants. At least one Diet group is different from the rest (permanova, *p*=2×10^-4^).

### Diversity of specific food groups can be calculated from NHANES data

Out of the 3,641 individuals, 50% (n=1,822) reported consuming at least one nuts/seeds/legumes food item during the two days of NHANES records. They reported consuming a total of 3,460 nuts/seeds/legumes food items, and there were 237 unique foods. The alpha-diversity index ranged from 0 to 1.95 (**Table 2**). We defined four dietary intake groups based on the Shannon’s diversity in the different nuts, seeds, and legumes consumed over two days: DivNA (n=1,819), Div0 (n=1,105), Div1 (n=360), and Div2 (n=357) (**Table 2**).

The ratios of males and females in each DivGroup were not different, but the other sociodemographic features, i.e., age, race/ethnicity, income, and education were different among the DivGroups (chi-square test, **Table 3**). The proportion of Asians increased from 6% to 23% as the diversity in nuts/seeds/legumes consumption increased. Notably, 29% of DivNA individuals, who did not consume nuts/seeds/legumes, had Family IPR ≥ 3.00; whereas 50% of Div2 group individuals had Family IPR ≥ 3.00. For education, 20% of the DivNA individuals had a college degree or higher, whereas 47% of Div2 group individuals did.

### Food group diversity is a marker of dietary quality

Diets of the diversity index groups differed in many of the food groups and nutrients (**Table 4**). The DivGroups with high nuts/seeds/legumes were generally eating more legumes and had higher percentages of legumes in protein foods than the low diversity groups, as expected. Interestingly, the higher the nuts/seeds/legumes diversity, the more vegetables and fruits and total energy were consumed (*p*-ANOVA <0.0001 for all three). Because of that, the nutrient intake was adjusted per 2,000 kcal. After adjustment, carbohydrate intake was not different among the DivGroups (*p*-ANOVA=0.476), but protein, dietary fiber, total fat, saturated fat, total unsaturated fat, and added sugar intakes were different such that the more diverse nuts/seeds/legumes consumption coincided with more dietary fiber, total fat, total unsaturated fat, and less total saturated fat and added sugars consumption. Interestingly, alcohol consumption was similar among the DivGroups (*p*-ANOVA=0.095).

### Legume diversity is associated with lower waist circumference

We modeled the association between waist circumference and nuts/seeds/legumes diversity, incorporating covariates (age, gender, ethnicity, income, education, and energy intake). ANCOVA indicated all the terms were significantly associated with waist circumference, except income and energy (*p*=0.238 and *p*=0.102, respectively; **Table 5**). Waist circumferences were different among the DivGroups (*p* <0.01, **Table 5**), and the group that did not consume nuts/seeds/legumes had 3.8 cm larger waist circumference than those who consumed the most diverse nuts/seeds/legumes foods (*p* <0.001, Tukey-adjusted, **Table 6**). In addition, the Div0 group that consumed only one nuts/seeds/legumes food had a 3.4 cm larger waist circumference than Div2 (*p* <0.01, Tukey-adjusted, **Table 6**). Similar results were found for BMI (see supplemental).

**Table 5.** ANCOVA table for Waist circumference.

|  | Sum Sq | Df | *F*-value | *p*-value |
| --- | --- | --- | --- | --- |
| (Intercept) | 968293 | 1 | 3684 | <0.0001 |
| DivGroup | 3963 | 3 | 5 | <0.01 |
| Age | 37101 | 2 | 71 | <0.0001 |
| Gender | 5027 | 1 | 19 | <0.0001 |
| Ethnicity | 31716 | 4 | 30 | <0.0001 |
| Family IPR | 755 | 2 | 1 | 0.238 |
| Education | 4204 | 2 | 8 | <0.001 |
| KCAL | 701 | 1 | 3 | 0.102 |
| Residual | 952704 | 3625 | . | . |

Df: degrees of freedom.

**Table 6.** Emmeans and contrast of waist circumference (cm) of the four levels of DivGroup. SE: standard error; CL: confidence limits.

| DivGroup | Contrast | Emmean | SE | Lower CL | Upper CL | *p*-value |
| --- | --- | --- | --- | --- | --- | --- |
| DivNA | . | 99.8 | 0.5 | 98.4 | 101.2 | . |
| Div0 | . | 99.5 | 0.6 | 97.8 | 101.1 | . |
| Div1 | . | 99.3 | 0.9 | 96.7 | 101.8 | . |
| Div2 | . | 96.1 | 0.9 | 93.4 | 98.7 | . |
| . | DivNA - Div0 | 0.4 | 0.6 | -1.4 | 2.1 | 0.942 |
| . | DivNA - Div1 | 0.5 | 1 | -2.2 | 3.2 | 0.943 |
| . | DivNA - Div2 | 3.8 | 1 | 1 | 6.5 | <0.001 |
| . | Div0 - Div1 | 0.2 | 1 | -2.6 | 3 | 0.998 |
| . | Div0 - Div2 | 3.4 | 1 | 0.6 | 6.2 | <0.01 |
| . | Div1 - Div2 | 3.2 | 1.2 | -0.2 | 6.6 | 0.04 |

## Discussion

The visualized proportion of energy from macronutrients and foodtrees, and the separation of individuals on five various diets on a PCoA plane demonstrates the utility of DietR to analyze dietary data collected on ASA24 or NHANES with general summarization, clustering analyses, incorporation of hierarchical grouping of food items, and ordination analyses based on the tree-based information.

Furthermore, DietR provides researchers with a tool to calculate dietary diversity for overall diet and within specific food groups or categories. The importance of diet diversity for adequate nutrition intake has long been discussed in the literature, and dietary diversity indices have been used to assess micronutrient adequacy, especially in developing countries (14). However, the relationship between dietary diversity indices and health outcomes such as non-communicable diseases is not straightforward (15). Thus, the diversity of specific food types instead of all foods consumed could be examined further for those food types that are likely to have positive effects on health. Here, we preliminarily examined nuts, seeds, and legumes diversity and demonstrated it as a potentially useful index to explore the health-promoting effects of nuts/seeds/legumes consumption (16).

We defined four dietary intake groups based on legume alpha-diversity (Shannon’s diversity, intake of multiple different nuts, seeds, and legumes consumed over 2 days) for n=3,641. The highest legume alpha-diversity group, Div2, had a lower waist circumference and a lower BMI, after adjustment for age, gender, ethnicity, and education variables. This association encourages more studies to be conducted to examine the hypothesis that the consumption of diverse nuts/seeds/legumes foods may contribute to a lowered risk of developing cardio-metabolic diseases, as a reduction in waist circumference is important to the prevention of cardio-metabolic diseases (17). In addition, the Div2 group had healthier dietary patterns that are high in legumes, total fruit, and vegetables, the consumption of which are encouraged by Dietary Guidelines for Americans (18). Div2 group also had less saturated fat and added sugar, and higher unsaturated fat intake, suggesting that the diversity in nuts/seeds/legumes consumption resulted in separation of individuals who are following healthy diets.

Taken together, the findings support the DietR approach, which is to apply ecological metrics to dietary data to calculate dietary diversity and to generate diversity of food categories that behave in expected ways. The DietR package provides a set of streamlined R functions to clean, analyze, and visualize dietary data from ASA24 and NHANES. By performing all the procedures in R, users can produce consistent and customized outputs with the flexibility that R, an open source tool with global users, offers.

We caution any causal or etiological interpretation of the data presented here. Our goal is to demonstrate the capabilities of DietR with an example analysis. We recognize that the results are presented without full incorporation of potential confounders. Exercise levels, for example, could be related to waist circumference and BMI as well as diversity in nuts/seeds/legumes consumption, as those who have higher healthy diet scores tend to be health-conscious and to follow regular exercise routines (19,20). Further investigation could be carried out with a larger sample size, using not only physical activity levels, but also smoking and drinking habits as potential covariates.

In conclusion, DietR offers functions to analyze dietary data collected on ASA24 or NHANES including data-cleaning, clustering analyses, and tree-based analyses. Moreover, DietR provides users with tools to calculate diversity metrics of specific food groups. New dietary diversity metrics it can produce are useful and could reveal interesting relationships between food and health in future studies.

## Data Availability

DietR package, including the ASA24 demonstration data, is available for download at https://github.com/computational-nutrition-lab/DietR.

https://computational-nutrition-lab.github.io/DietR/

https://www.n.cdc.gov/nchs/nhanes/continuousnhanes/default.aspx?BeginYear=2015

## Abbreviations

ANCOVA: Analysis of Covariance
ANOVA: Analysis of Variance
ASA24: Automated Self-Administered 24-hour dietary assessment tool
BMI: Body Mass Index
FNDDS: Food and Nutrient Database for Dietary Studies
IFC: Individual Food Consumption Table
IPR: Income to Poverty Ratio
NHANES: National Health and Nutrition Examination Survey
PCA: Principal Component Analysis
PCoA: Principal Coordinate Analysis

## Acknowledgements

The authors would like to thank Mo Hutti for the create_corr_frame function which generates a correlation table with ordination axes and variables; Pajau Vangay for the collapse_by_correlation function which removes correlated variables; and Suzie Hoops for matrix multiplication operation and insights into statistical analyses.

## Authors’ contributions to manuscript

RS and AJJ conceptualized and created the DietR package and wrote the manuscript. RS analyzed data. RS and AJJ had primary responsibility for final content. MAP and DJ were involved in the development of statistical models and data interpretation, and critically revised the manuscript. All authors read and approved the final manuscript.

## Software and data availability

The tutorial can be accessed on https://computational-nutrition-lab.github.io/DietR/.

The National Health And Nutrition Survey data can be accessed at https://www.n.cdc.gov/nchs/nhanes/continuousnhanes/default.aspx?BeginYear=2015.

