## Supplemental methods and tables for "Dietary pattern and diversity analysis using ‘DietR’ package in R"

### Supplementary Materials

**Supplementary methods**

Automated Self-Administered 24-hour dietary assessment tool (ASA24) is a web-based dietary study platform where researchers set up a study, and recruited participants enter their dietary records into their own page accessed by participant-specific username and password (1). The researcher decides the duration of each study and additional questions to ask regarding potential predictors of health as metadata. Collected data can be downloaded by bulk during or after data collection by the researcher. Downloaded data consists of six comma-separated values (CSV) files.

One of the CSV files, Items.csv, has all individual food items and related information entered by each participant for each day and eating occasion. Total.csv has summed values of the items reported for each day for each participant. However, Items.csv file should be inspected first, and if modifications are made, the total should be re-calculated with the inspected (QC-ed) Items data.

**Difference in BMI by DivGroup**

For BMI, the initial model with all the covariates indicated that income and energy did not have an effect ($p$=0.462 and $p$=0.202, respectively); thus, income and energy were removed from the model, and ANCOVA was run again with the rest of the covariates (**Supplementary Table 2**). BMI was different among the DivGroups ($p$<0.01, **Supplementary Table2**), and the group that did not consume nuts/seeds/legumes had 1.4 larger BMI than those who consumed the most diverse nuts/seeds/legumes foods ($p$<0.01,**Supplementary Table 3**). In addition, the Div0 group that consumed only one nuts/seeds/legumes food had a 1.4 larger BMI than Div2 ($p$<0.01, **Supplementary Table 3**). The other contrasts were not different.

**Supplementary Table 1** Dependency packages of DietR.

| Package | Use in the DietR package. | Reference |
|---|---|---|
| data.tree | Build foodtrees. | (2) |
| factoextra | Perform Silhouette and the Gap methods to find an optimal k in k-means analysis. | (3) |
| ggfortify | Generate PCA biplots. | (4) |
| ggplot2 | Generate various charts including barplots and scatterplots. | (5) |
| ggtree | Visualize foodtrees. | (6) |
| gridExtra | Generate multiple-panel charts. | (7) |
| pairwiseAdonis | Perform pairwise permanova test for groups. | (8) |
| phyloseq | Build a phyloseq object, perform ordination using the vegan package. | (9) |
| reshape2 | Convert short tables to long tables. | (10) |
| SASxport | Import .XPT files downloaded from the NHANES website and convert the data into a dataframe in R. | (11) |
| vegan | Calculate distances between participants, perform ordination and permanova | (12) |

**Supplementary Table 2** ANCOVA table of BMI. Df: degrees of freedom.

| | Sum Sq | Df | *F*-value | *p*-value |
|---|---|---|---|---|
| (Intercept) | 75713 | 1 | 1620 | <0.0001 |
| DivGroup | 576 | 3 | 4 | <0.01 |
| Age | 1134 | 2 | 12 | <0.0001 |
| Gender | 631 | 1 | 13 | <0.001 |
| Ethnicity | 5994 | 4 | 32 | <0.0001 |
| Family IPR | 72 | 2 | 1 | 0.462 |
| Education | 733 | 2 | 8 | <0.001 |
| KCAL | 76 | 1 | 2 | 0.202 |
| Residuals | 169456 | 3625 | . | . |

**Supplementary Table 3** Emmeans and contrast of BMI of the four levels of DivGroup.SE: standard error; CL: confidence limits.

| DivGroup | Contrast | Emmean | SE | Lower CL | Upper CL | *p*-value |
|---|---|---|---|---|---|---|
| DivNA | . | 29.2 | 0.2 | 28.6 | 29.8 | . |
| Div0 | . | 29.2 | 0.2 | 28.6 | 29.9 | . |
| Div1 | . | 29.0 | 0.4 | 28 | 30.1 | . |
| Div2 | . | 27.8 | 0.4 | 26.7 | 28.9 | . |
| . | DivNA - Div0 | 0 | 0.3 | -0.8 | 0.7 | 1 |
| . | DivNA - Div1 | 0.2 | 0.4 | -0.9 | 1.3 | 0.963 |
| . | DivNA - Div2 | 1.4 | 0.4 | 0.2 | 2.5 | <0.01 |
| . | Div0 - Div1 | 0.2 | 0.4 | -1 | 1.4 | 0.962 |
| . | Div0 - Div2 | 1.4 | 0.4 | 0.2 | 2.6 | <0.01 |
| . | Div1 - Div2 | 1.2 | 0.5 | -0.2 | 2.6 | 0.092 |

### Sample code

DietR and R code to load ASA24 items data set, compute total food consumption per participant, add participant's metadata to totals, and review and omit potential outlier records.

```
# Import source code to run the analyses to follow.
  source("lib/specify_data_dir.R")
  source("lib/load_clean_ASA24.R")
  source("lib/average.by.R")
  source("lib/QCOutliers.R")
  source("lib/Food_tree_scripts/format.foods_2.r")

# You can come back to the main directory by:
  setwd(main_wd)

# ==================================================================================
# Load ASA24 data
# ==================================================================================

# Specify the directory where the data is.
  SpecifyDataDirectory(directory.name= "eg_data/VVKAJ/")

# Load your unprocessed (raw) food items-level data (as downloaded from the ASA24
study website).
# The csv file will be loaded as a dataframe in R and be named as items_raw.
  items_raw <- read.csv("Raw_data/VVKAJ_Items.csv", sep = ",", header=T)

# items_raw has a column called "Food_Description", but this needs to be changed to
"Main.food.description". Change the column name.
  names(items_raw)[names(items_raw) == "Food_Description"] <- "Main.food.description"

# Check if any column names match with "Main.food.description". If there is a match,
it will be printed.
  names(items_raw)[names(items_raw) == "Main.food.description"]

  # [Note] The numbers in the square brackets of the output indicate the sequential
number of each element to help count the number of elements.

# Save the items file as a .txt file. This command saves the object "items_raw" as
a .txt file with the specified filename using the write.table function.
  write.table(items_raw, "VVKAJ_Items.txt", sep="\t", row.names=F)

# Special characters common in food names in dietary data such as "'", ",", "%" may
interfere correct data loading in R; thus, we replace them with an underscore "_".

# Format foods so that special characters will be replaced with "_". "_f" stands for
"formatted".
```

```
  FormatFoods(input_fn =  "VVKAJ_Items.txt",
             output_fn = "VVKAJ_Items_f.txt")

# [Note] It is best practice to avoid overwriting your raw data. Always save
formatted/manipulated versions as a new file as described above.

# Load the Items_f.txt file to take a look at it.
# You need the "quote=""" and "colClasses="character"" arguments to ignore quotation
marks (do not regard them as a cell separator) and to load all the columns as
characters so that FoodID will keep the trailing ".0".
  items_f <- read.delim("VVKAJ_Items_f.txt", quote="", colClasses="character")

# Add a human-readable sample identifier (SampleID) with a desired prefix, and save
it as a txt file. SampleIDs are IDs unique to each combination of users and day and
represent days of dietary intake in this dataset.
  AddSampleIDtoItems(input.fn="VVKAJ_Items_f.txt", user.name="UserName",
recall.no="RecallNo", prefix="vvkaj.", out.fn="VVKAJ_Items_f_id.txt")

# Load the formatted Items file with SampleID added.
  items_f_id <- read.delim("VVKAJ_Items_f_id.txt", quote="", colClasses="character")

# ================================================================================
#  Merge individuals' metadata to items.
# ================================================================================

# ind_metadata has the participants' gender, age, height, weight, BMI, and
Waist.Circumference, etc.
# If desired, this individual-specific information can be added to items data.

# Load ind_metadata.txt.
  ind_metadata <- read.table("ind_metadata.txt", sep="\t", header=T)

# Add this metadata of each participant to totals or items.
# 'NA' will be inserted to UserNames which are not in ind_metadata.
  items_f_id_s_m <- merge(x=items_f_id_s, y=ind_metadata, by="UserName", all.x=T)

# Save the merged dataframe as a .txt file.
  write.table(items_f_id_s_m, "VVKAJ_Items_f_id_s_m.txt", sep="\t", row.names=F,
quote=F)

# ================================================================================
# Generate new totals file from the items file.
# ================================================================================

# Use one of the input files saved above as an input for calculating totals.
# Specify which columns have usernames and Recall.No., which has the recorded days.
  GenerateTotals(inputfn =   "VVKAJ_Items_f_id_s_m.txt",
                User.Name = "UserName",
```

```
                 Recall.No = "RecallNo",
                 outfn =     "VVKAJ_Tot.txt")

# Load the total file generated above.
  new_totals <- read.table("VVKAJ_Tot.txt", header=T, sep="\t")

# ================================================================================
#  Add the participants' metadata back to totals.
# ================================================================================

# Load ind_metadata.txt if you have not done so.
  ind_metadata <- read.table("ind_metadata.txt", sep="\t", header=T)

# Add this metadata of each participant to totals.
# 'NA' will be inserted to UserNames which are not in ind_metadata.
  new_totals_m <- merge(x=new_totals, y=ind_metadata, by="UserName", all.x=T)

# Save the merged dataframe as a .txt file.
  write.table(new_totals_m, "VVKAJ_Tot_m.txt", sep="\t", row.names=F, quote=F)

# ================================================================================
# Calculate the mean of totals/participant
# ================================================================================

# Calculate the mean of the totals data across all the days for each participant.
  AverageBy(data= new_totals, by= "UserName", start.col= "FoodAmt", end.col=
"A_DRINKS", outfn="VVKAJ_Tot_mean.txt")

# Load the output for further processing.
  new_totals_mean <- read.table("VVKAJ_Tot_mean.txt", header=T, sep="\t")

# ================================================================================
# Add the participants' metadata to the mean totals.
# ================================================================================

# Load ind_metadata.txt if you have not done so.
  ind_metadata <- read.table("ind_metadata.txt", sep="\t", header=T)

# Add this metadata of each participant in the mean totals.
# 'NA' will be inserted to UserNames which are not in ind_metadata.
  new_totals_mean_m <- merge(x=new_totals_mean, y=ind_metadata, by="UserName",
all.x=T)

# Save the merged dataframe as a .txt file.
  write.table(new_totals_mean_m, "VVKAJ_Tot_mean_m.txt", sep="\t", row.names=F,
quote=F)

# ================================================================================
```

```
# Quality Control (QC) for the mean totals data
# ==================================================================================

# Split your dataset to males and females because different thresholds apply for
males and females.
  new_totals_mean_m_M <- subset(new_totals_mean_m, Gender=="M")
  new_totals_mean_m_F <- subset(new_totals_mean_m, Gender=="F")

# ---------------------------------------------------------------------------------
# QC for males
# Define your males totals dataset to be used as input.
  QCtotals <- new_totals_mean_m_M

# Flag if KCAL is <650 or >5700 --> ask remove or not --> if yes, remove those rows
  QCOutliers(input.data = QCtotals, target.colname = "KCAL", min = 650, max = 5700)

# If you find potential outlier(s) here, click "No", and view those
# total(s) with their other nutrient intake information by running the following;
  KCAL_outliers <- subset(QCtotals, KCAL < 650 | KCAL > 5700)

  # Sort the rows by KCAL and show only the specified variables.
  KCAL_outliers[order(KCAL_outliers$KCAL, decreasing = T),
              c('UserName', 'KCAL', 'FoodAmt', 'PROT', 'TFAT', 'CARB')]

# If you think it is a true outlier, then run the QCOutliers command for KCAL again,
and click "Yes" to remove the outlier. Here for this tutorial, we will remove this
individual.
  QCOutliers(input.data = QCtotals, target.colname = "KCAL", min = 650, max = 5700)

# Continue the QC process with other variables.

# Flag if PROT is <25 or >240 --> ask remove or not --> if yes, remove those rows
  QCOutliers(input.data = QCtotals, target.colname = "PROT", min = 25, max = 240)

# Flag if TFAT is <25 or >230 --> ask remove or not --> if yes, remove those rows
  QCOutliers(input.data = QCtotals, target.colname = "TFAT", min = 25, max = 230)

# Flag if VC (Vitamin C) is <5 or >400 --> ask remove or not --> if yes, remove those
rows
  QCOutliers(input.data = QCtotals, target.colname = "VC", min = 5, max = 400)

# Name the males totals after QC.
  QCed_M <- QCtotals

# ---------------------------------------------------------------------------------
# QC for females
  # Define your females totals dataset to be used as input.
  QCtotals <- new_totals_mean_m_F
```

```
  # Flag if KCAL is <600 or >4400 --> ask remove or not --> if yes, remove those rows
  QCOutliers(input.data = QCtotals, target.colname = "KCAL", min = 600, max = 4400)

  # Flag if PROT is <10 or >180 --> ask remove or not --> if yes, remove those rows
  QCOutliers(input.data = QCtotals, target.colname = "PROT", min = 10, max = 180)

  # Flag if TFAT is <15 or >185 --> ask remove or not --> if yes, remove those rows
  QCOutliers(input.data = QCtotals, target.colname = "TFAT", min = 15, max = 185)

  # Flag if VC (Vitamin C) is <5 or >350 --> ask remove or not --> if yes, remove
those rows
  QCOutliers(input.data = QCtotals, target.colname = "VC", min = 5, max = 350)

# Name the females totals after QC.
  QCed_F <- QCtotals

# ---------------------------------------------------------------------------------
# Combine the rows of M and F.
  QCtotals_MF <- rbind(QCed_M, QCed_F)

# Save as a .txt file.
  write.table(QCtotals_MF, "VVKAJ_Tot_mean_m_QCed.txt", sep="\t", quote=F,
row.names=F)
```
